# EVALUATION OF A METAGENOMIC NEXT-GENERATION SEQUENCING ASSAY WITH A NOVEL HOST DEPLETION METHOD FOR PATHOGEN IDENTIFICATION IN SEPTIC PATIENTS

**DOI:** 10.1101/2023.03.28.23287867

**Authors:** Yen-Chia Chen, Po-Hsiang Liao, Yen-Wen Chen, Chia-Ming Chang, Maurice Chan, Deng Fong Chao, Yizhen Lin, Jiahao Chang, Hau Hung, Mengchu Wu, David Hung-Tsang Yen

## Abstract

**Background:** The traditional diagnosis of sepsis has always been based on microbial blood culture (BC). However, BC suffers from (1) long culture cycle, leading to delay in results, and (2) low diagnostic yields. Metagenomic next-generation sequencing (mNGS) has been proposed as an efficient and agnostic option that potentially overcomes these issues. In this study, a mNGS workflow utilizing a novel filter to specifically capture white blood cells and deplete host DNA background, was evaluated against BC results, as well as mNGS without host depletion, for pathogen identification.

**Materials and Methods:** Patients admitted to Taipei Veterans General Hospital (TVGH) with suspected sepsis were recruited to the study approved by the IRB. Blood sample was taken for BC (designated as BC1) before any antibiotic exposure. Upon patient enrolment, blood was taken again and divided in 3 portions with one used for the 2^nd^ BC (BC2). The other two were used for mNGS with one processed with the filter and the other without filtering, to assess the effectiveness of host-depletion by the filter.

**Results:** A total of 50 patients were recruited among which 45 had results for all 4 tests. mNGS with filter had the highest positive rate of 74.4%, followed by mNGS without filter and BC1 (51.1% and 50.0% respectively), while the 2^nd^ BC had the lowest positive rate of 22.0%. Further, mNGS was less sensitive to antibiotics exposure as compared to BC. The overall correlation between samples with vs without filtration (R^2^=0.96) confirmed that filtration does not affect microbial composition in a sample. For the BC positive samples, the effect of host depletion by filtration increased microbial target reads/million QC reads from 46 reads to 243 reads on average. Microbial reads enrichment by the filter appeared to be more effective for the samples with lower microbial concentration, thus increasing the test sensitivity over mNGS without filter. Using the 2^nd^ BC results as reference, mNGS with filter and mNGS without filter exhibited sensitivities of 81.8% and 63.6%.

**Conclusion:** The mNGS with filter was able to recover most of the pathogens identified by clinical BC and achieved the highest diagnostic yield. With the clinical implementation to complete the workflow within 24 hours, it has the potential to overcome slow turnaround and low diagnostic yield issues of traditional BC.

## Introduction

Sepsis is a life-threatening condition arising from the human immune response to the infection of the bloodstream [1]. Sepsis can progress into septic shock, organ failure and death if diagnosis and correct treatment are not timely made [2]. Morbidity and mortality rates of blood stream infections (BSIs) are especially high in the intensive care units (ICU) and neonatology units of hospitals [3]. The traditional workflow to diagnose sepsis in clinical microbiology laboratories starts with blood culturing (BC), necessitating between 1 ml and 10 ml of blood per BC bottle, often in automated systems such as the BACTECTM (Becton Dickinson-BD, Maryland, USA) or BacT/Alert (bioMérieux, Marcy l’Etoile, France. A positive BC is followed by Gram staining. Bacterial genus and species identification can be performed on the cultured strain with biochemical assays or MALDI-TOF mass spectrometry (MS). Antibiotics susceptibility testing of the cultured strain is generally performed with microbiological assays.

The short-comings of traditional methods have been expounded [4]: they do not detect viruses and parasites; some bacterial pathogens are difficult to culture; the culture cycle is long, so the results are delayed; and it is difficult to identify mixed infections (due to single colony picking). One of the major shortcomings of traditional methods is its low diagnostic yield (positive rates). In one study [4], the positivity rates of blood, sputum, and BALF using traditional detection methods were 14% (13/90), 38% (34/90), 22% (10/45), respectively. In immunocompromised and immunocompetent groups of patients, 47% (14/30) and 27% (16/60) of patients, respectively, were positive by NGS but negative by traditional detection methods.

Despite the availability of rapid molecular assays, most early antibiotic treatment is still empirical. According to one study [5], approximately 46% of early empirical antibiotic treatment was inappropriate, directly leading to a sepsis-related mortality rate of almost 35%. Approximately 50% of antibiotics administered are unnecessary or too broad-spectrum, which increases the toxicity of drugs and the incidence of bacterial resistance [5]. Therefore, early identification of pathogens is particularly important to enable targeted antibiotic therapy. Several reports have presented the potential advantages of molecular diagnostics over BC, such as a shorter turnaround time, detection of difficult to grow bacteria or detection after prior intake of antibiotics which inhibits bacterial growth [6]. Caliendo et al. reported that molecular diagnosis of BSI can reduce hospitalization rates and length of ICU stays, and decrease mortality due to BSI [7].

In recent years, metagenomic NGS has been proposed as a more efficient and accurate means for pathogen diagnosis and expanded the options of diagnostic strategies for pathogen identification [4] [8] [9] [10]. mNGS is based on the “agnostic” extraction and sequencing of all nucleic acids in a patient’s specimen in parallel, so as to obtain the sequence of host and microorganisms. mNGS can be applied to a wide range of specimen types (sputum, throat swab, blood, alveolar lavage fluid, pleural fluid, cerebrospinal fluid, pus, tissue specimens, etc), and it can directly detect nucleic acids in clinical samples without biasness or selectivity. Viruses, bacteria, fungi, and parasites can be detected in an unbiased manner. mNGS could detect and identify multiple pathogens simultaneously.

NGS was used for the clinical diagnosis of neuroleptospirosis in a 14-year-old critically ill boy with meningoencephalitis; this case was the first to demonstrate the utility of metagenomic NGS (mNGS) in providing clinically actionable information, as successful diagnosis prompted appropriate targeted antibiotic treatment and eventual recovery of the patient [11]. Huang et al. showed that in patients with pulmonary infection, the positive-rate of mNGS (88.30%) for pathogen detection in pulmonary infection was much higher than that of traditional detection methods (25.73%), while the specificity of mNGS (81.16%) was slightly lower than that of traditional detection methods (88.41%) [9]. Cheng and Yu [4] showed that, in a cohort of immunocompromised and immunocompetent sepsis patients, 77 (86%) were positive for 1 or more pathogens using NGS, and 50 (56%) were positive using traditional detection methods.

One widely recognised major hurdle in mNGS of blood samples is the overwhelming presence of human DNA, leading to vast wastage of sequencing real estate [11]. A few methods have been proposed for removing host DNA background, including differential lysis of human cells [12] and methylated human DNA removal [13]. Recently, host cell depletion device such as Devin fractionation filter has been made available for removing human nucleated cells. Incorporating such device into mNGS workflow imposes an efficient way to enrich microorganisms in the sample, to increase the sensitivity of the assay or decrease the total cost of the assay. The objective of this study is to evaluate the efficacy of the Devin fractionation filter in host depletion and enrichment of microbial genomic sequences in the mNGS workflow used in this study. Another objective of this study is to evaluate the clinical performance of the mNGS workflow incorporating Devin filtration, using a cohort of patients with suspected sepsis who had been admitted to Taipei Veterans General Hospital (TVGH).

## MATERIALS AND METHODS

### Patient samples

Blood samples were recruited from patients admitted to Emergency Department of Taipei Veterans General Hospital (TVGH) from April 2021 to September 2022 who were suspected of sepsis. The study has been approved by the Institutional Review Board (Ethics Committee) of TVGH (IRB number, 2021-03-013AC, approval date: March 22, 2021). As illustrated in Fig. 1, Blood sample for routine clinical blood culture was taken upon admission as the 1st blood culture (designated as BC1) which may be followed by antibiotic exposure for most of the cases. Blood sample for this study was then taken after consent was obtained. It was divided in 2 portions. One portion was used for blood culture designated as 2^nd^ blood culture (BC2) while the other portion of approximately 8 mL was used for mNGS.

**Fig. 1.**
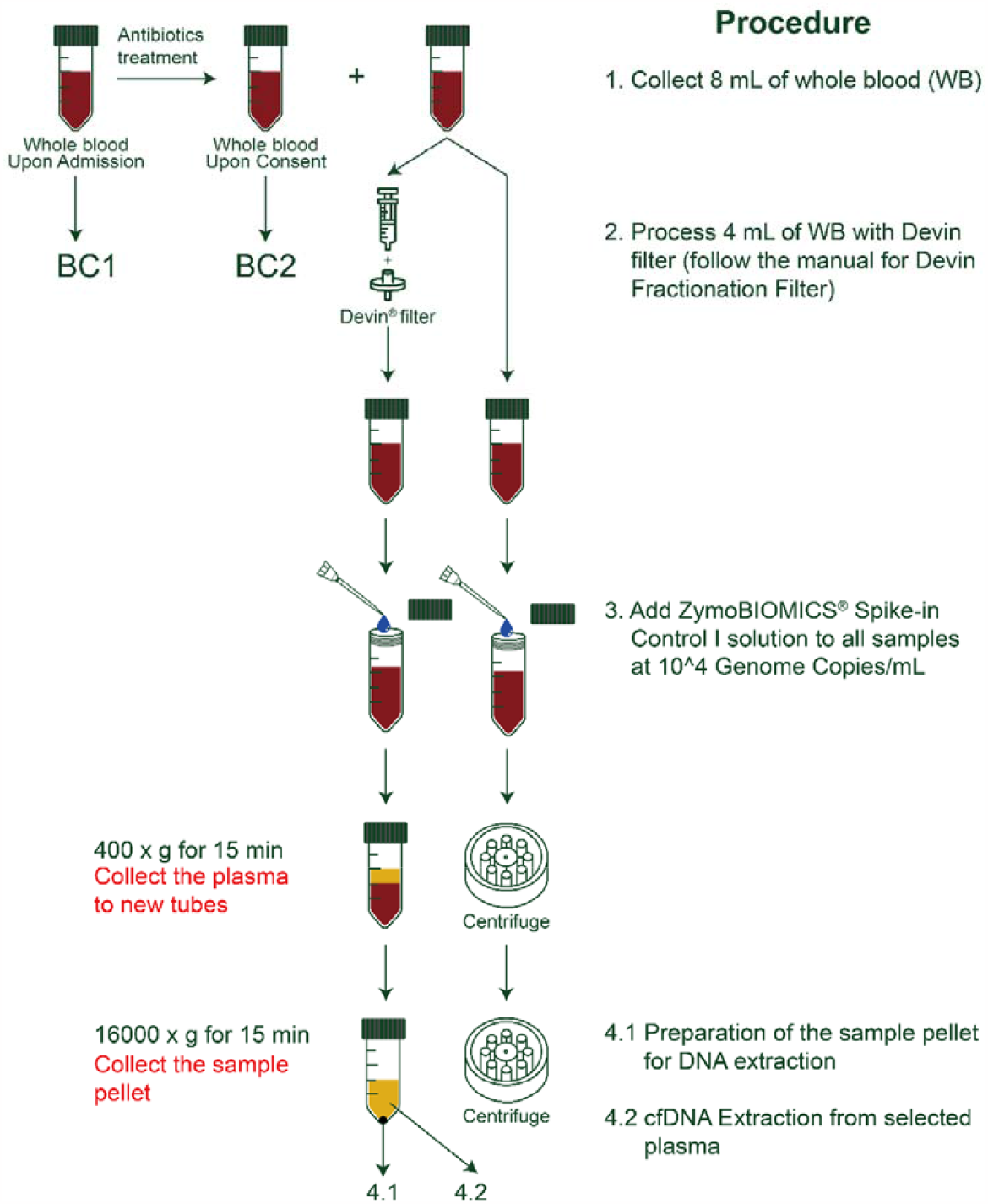
Experimental plan for this study.

### Host depletion using Devin™ fractionation filter

In order to assess the effectiveness of host-depletion by the Devin™ filter, the ∼8 mL whole blood sample was further divided into two portions, one processed for host-depletion using the Devin™ fraction filter (Micronbrane Medical, Taiwan), and the other processed without filtering. The Devin™ fractionation syringe filter series exploits the antifouling Zwitterionic coating technology to specifically capture the white blood cells (>99%) in the whole blood, while allowing other blood components to flow through the membrane. The filter was securely attached to a syringe on the one side. Approximately 4 mL of whole blood sample was transferred into the syringe. The syringe plunger was gently pressed down to push the sample through the filter to a 15 mL falcon tube. Both the filtered and unfiltered blood samples were used for the downstream assays.

### Sample processing and DNA extraction

As illustrated in Fig. 1, ZymoBIOMICS Spike-in Control I (High Microbial Load) (Zymo Research, Irvine, CA) which included two extremophiles bacterial species (*Imtechella halotolerans* and *Allobacillus halotolerans*) was added to all samples including NTC (No Template Control) at the concentration of 10^4 Genome Copies/mL to act as internal reference controls. Both the filtered and unfiltered blood samples were centrifuged at low speed (400g) for 15 min at room temperature to obtain the plasma. The plasma was then centrifuged at high speed (16,000g) to obtain the sample pellet for DNA extraction using Devin™ Microbial DNA Enrichment kit (Micronbrane Medical, Taiwan). For selected samples, the supernatant after the high-speed centrifugation was also obtained for cell-free DNA (cfDNA) extraction using iCatcher® Circulating cfDNA1000 Kit (CatchGene Co., Ltd., Taiwan).

### Library construction and NGS

Library preparation with DNA extracted from the pellet was carried out using Illumina DNA Prep Library Kit (Illumina, USA), according to manufacturer’s instructions. After DNA tagmentation, PCR amplification was carried out using the following conditions: (Top lid at 100°C); 68°C, 3min; 98°C, 3min; 15 cycles of 98°C, 45sec/ 62°C, 30sec/ 68°C, 2min; 68°C, 1min; 10° C, ∞. After amplification, the reaction was purified using Sample Purification Beads and eluted with 21 µl of Resuspension Buffer (Illumina, USA). 20 µl supernatant was transferred to a new Lobind Eppendorf tube and stored at -20°C. The constructed libraries were sent to a service provider for sequencing on Illumina NovaSeq platform to obtain a minimum of 20 million reads per sample at 150bp. NGS Library preparation for cfDNA from plasma were carried out using xGen™ ssDNA & Low-Input DNA Library Preparation Kit (Integrated DNA Technologies, USA) following manufacturer’s protocol.

### Bioinformatics pipeline and results interpretation

All sequencing reads were trimmed for adapter sequences and poor-quality bases (<Q30) using fastp v0.23.2 [14]. Reads mapped to human genome (GRCh38) were performed with bwa 0.7.17 (bwa-mem algorithm) [15]. The remaining reads were aligned to microbial database with bwa 0.7.17. A set of representative genomes for microorganisms (bacteria, viruses, fungi, protozoa, and other multicellular eukaryotic pathogens) from the NCBI Nucleotide and Genome databases was used for microbial alignment. The final database consisted of about 1400 genomes. For each sample, the % of each microorganism (microbe %) was calculated as the % of the classified reads in the total microbial reads. The absolute classified reads were normalized to one million QC reads named as RPM (Reads per Million) for each microorganism. A non-template control (NTC) was mandated for each reagent batch. The Microbe % and RPM was calculated in the same way for NTC as for the samples. The NTC was used to filter out contaminants from the laboratory and reagents. Sample to NTC ratios were computed using % of reads/total microbial reads (microbe %_SAMPLE_ : microbe %_NTC_). Microorganisms were kept only if their % were found ≥ 5-folds in samples than in controls and their RPM ≥ 5. mNGS results were primarily compared with microbial identification results from BC2 (the reference result), since both mNGS and BC2 resulted from the same blood draw. Subsequently, BC1 results as well as other culture results if available were also used for evaluating the overall performance of mNGS. Prior to finalization of read-length of analysis, a subset of 11 BC1 positive samples were subject to analysis based on 100bp, 120bp, and 150bp (Supplemental Table 1). Analysis using various read length did not affect the recovery of positive sequences. As such, 100bp-based analysis was selected to be used in this study as the sequencing and analysis time of 100bp read length is the most feasible condition to finish the whole metagenomic NGS workflow within 24 hours for real clinical application.

## RESULTS

### Clinical Performance of mNGS with and without host depletion

Total of 50 test subjects (identified by sample number in Table 1) were recruited for this study. These were patients admitted into Emergency Department with suspected sepsis. The microbial culture results from the first blood draw (BC1) were the routine clinical results. This may be followed by antibiotic exposure in most cases. Blood sample for this study (BC2) was then taken after consent was obtained. For testing the efficacy of host depletion, each sample was processed with and without Devin filtration followed by the same DNA extraction, NGS library construction process and sequenced at 150bp. Three cases (#25, #31 and #37) had library preparation failure for both with and without filter samples. Two cases (#27 and #38) failed library preparation failure for the without filter sample only. Hence, the success rate of library prep was 92% (92/100).

**Table 1:**
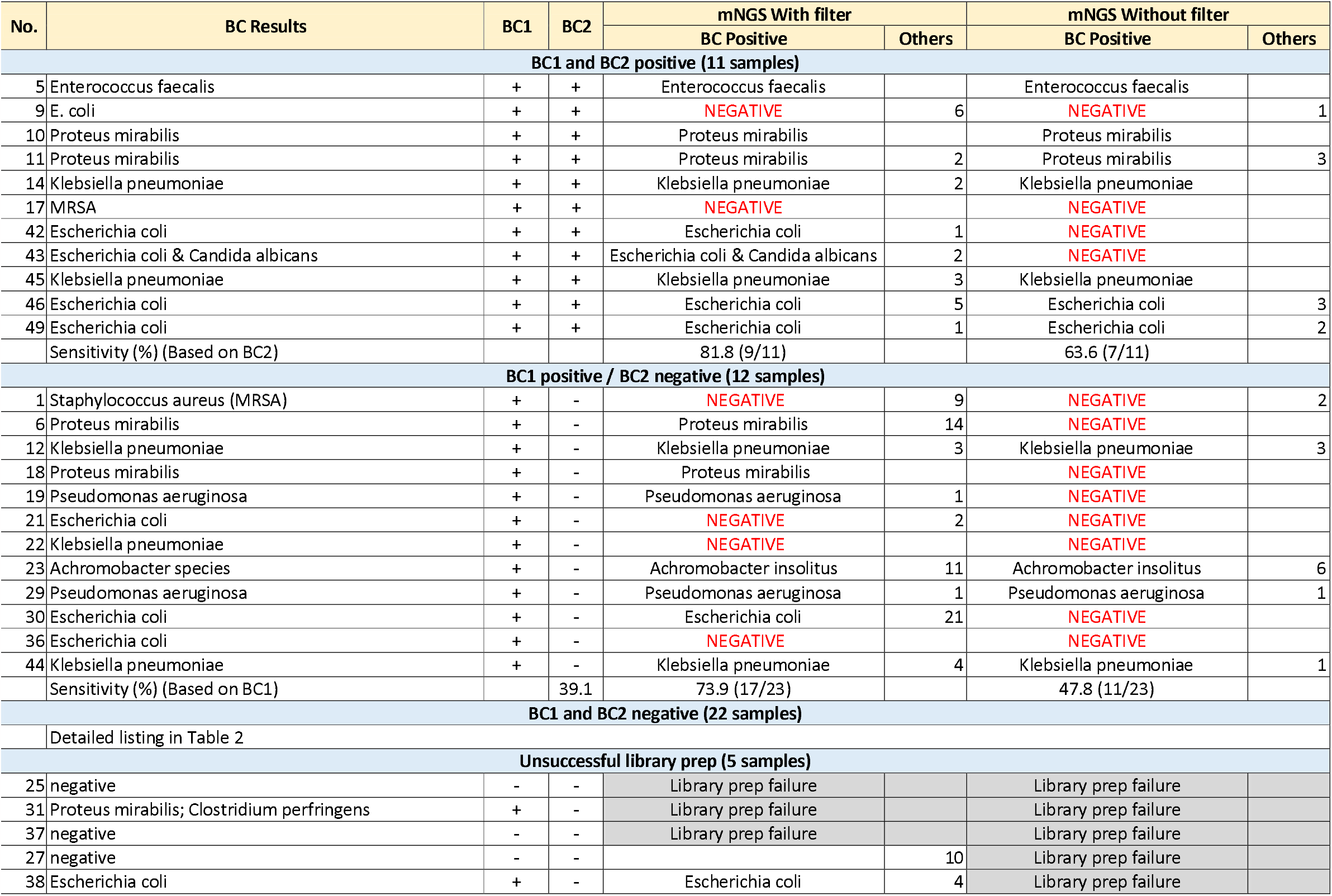
Summary of mNGS results correlated with clinical microbial culture and reference microbial culture results.

As the turn-around time (TAT) is one of the crucial parameters for pathogen detection method, we first evaluated the microorganism classification performance by different sequencing read lengths in the 11 samples having positive culture results for both BC1 and BC2. As shown in Supplementary Table 1, the sequencing analysis results were compared among different read length of 150, 120 and 100bp in both mNGS with and without Devin™ filter. Reads were normalized to million QC reads (RPM) for comparison among samples. Overall, a highly comparable results were observed for different read length among all 11 samples. Most importantly, when applying the tentative cut-off criteria for potential pathogen identification, viz fold change (microbe %_SAMPLE_ : microbe %_NTC_) ≥ 5 and microbial RPM ≥ 5, the results were unaffected by different read length. Hence, analysis with 100 bp read lengths offers the most efficient option, and was selected to be used in this study. The summary of the reads classification results at 100 bp for all samples was shown in the Supplementary Table 2.

Both fold change (microbe %_SAMPLE_ : microbe %_NTC_) and individual species RPM (Reads per Million) are direct indicators of the amount of species in the sample. Using BC results as a comparison, setting both indictors at a threshold of ≥ 5 for pathogen calling appeared to be reasonable criteria to achieve good sensitivity and specificity. The microorganisms identified passing these criteria for all samples were listed in Supplementary Table 3. The identification results of all samples were then compared to both the BC1 and BC2 results and summarized in Tables 1 and 2. As shown in Table 1, when comparing to BC2 positive results, mNGS with filtration was able to detect the pathogens consistent with the microbial culture in 9 out 11 samples while mNGS without out filter was able to detect 7 out of 11 samples. Hence the sensitivity of mNGS with or without filtration was 81.8% and 63.6% respectively. Filtration let to higher sensitivity performance through increasing overall number of microbial reads and target species reads in most of the samples. In sample #17, both mNGS methods failed to detect MRSA. In sample #9, both mNGS failed to identify *E. coli* but were able to detect other species passing the criteria set. Further, mNGS (with filtration) provided the highest diagnostic yield (35/47; 74.4%), followed by mNGS (without filtration) (23/45; 51.1%), BC1 (25/50; 50%) and BC2 (11/50; 22%). Out of 12 samples which were positive for BC1 but negative for BC2, mNGS detected pathogen in 8 (with filter) and 4 (without filter) samples consistent with the culture results of BC1. Hence, this data also suggested that mNGS is less sensitive to antibiotic treatment as compared to culture methods.

**Table 2:** Potential pathogens identified by mNGS in the 22 blood culture negative samples.

| No. | # Potential Pathogen Identified |  | Clinical diagnosis | Other culture results | Correlation with mNGS |
| --- | --- | --- | --- | --- | --- |
|  | mNGS W/ filter | mNGS W/O filter |  |  |  |
| 2 | 1 |  | *Clostridium difficile* | stool: *Clostridium difficile* |  |
| 3 | 8 | 2 | *Pseudomonas aeruginosa* | urine: *Pseudomonas aeruginosa* |  |
| 4 | 7 | 3 | *Escherichia coli* | urine: *Escherichia coli* |  |
| 7 | 8 | 1 | *Escherichia coli* | urine: *Escherichia coli* |  |
| 8 | 9 | 11 | undetermined | urine: Gram positive cocci in chain | *Streptococcus suis* |
| 13 | 2 | 1 | *Escherichia coli* | urine: *Escherichia coli* |  |
| 15 | 2 | 1 | *Klebsiella pneumoniae* | blood (in other hospital): *Klebsiella pneumoniae* |  |
| 16 | 9 |  | *Escherichia coli* | urine: *Escherichia coli* |  |
| 20 |  |  | *Clostridium difficile* | PCR (+): *Clostridium difficile* |  |
| 24 | 1 | 2 | undetermined | negative for urine and sputum |  |
| 26 | 3 | 2 | undetermined | negative for urine and sputum |  |
| 28 | 2 | 1 | *Klebsiella pneumoniae* | abscess: *Klebsiella pneumoniae* | *Klebsiella pneumoniae* |
| 32 | 1 |  | *Escherichia coli* | urine: *Escherichia coli* |  |
| 33 |  |  | undetermined | urine: 1. *Aerococcus urinae*; 2. Gram negative bacilli |  |
| 34 |  |  | undetermined | urine: *Citrobacter freundii*; sputum: *Acinetobacter baumannii* (CRAB) |  |
| 35 |  |  | undetermined | negative for urine and sputum | Negative for mNGS |
| 39 | 1 |  | *Candida* | urine: Yeast |  |
| 40 |  |  | *Escherichia coli* | urine: *Escherichia coli* |  |
| 41 | 1 |  | undetermined | negative for urine and sputum | Torque teno virus |
| 47 | 9 | 4 | *Candida* | endo aspirate: *Candida krusei* and *Candida glabrata* |  |
| 48 | 1 | 2 | *Klebsiella pneumoniae* | abscess: *Klebsiella pneumoniae* | *Klebsiella pneumoniae* |
| 50 | 1 | 1 | *Streptococcus agalactiae* | urine: *Streptococcus agalactiae* | *Streptococcus agalactiae* |

In general, mNGS detected more microorganisms per sample as compared to microbial culture (see columns labelled as “Others” in Table 1 and “# Potential Pathogens Identified” in Table 2). Microbial detection by mNGS from blood culture negative samples, where both BC1 and BC2 were negative, is shown in Table 2. Interestingly, amongst the 22 blood culture negative samples (both BC1 and BC2 negative), microbial detection in 5 samples were correlated to additional testing results – mNGS results in 4 samples included a pathogen that correlated with other clinical results; 1 sample was negative for mNGS and had negative results from all routine tests. This suggested that although mNGS detected more pathogens as compared to microbial culture, the additional pathogens called may be correlatable to other clinical results, and may be clinically useful.

### Effect of Devin filtration on number and ratio of microbial reads

Although human reads remained the predominant class of sequence from whole blood samples despite the employment of host cell depletion by Devin™ filter, total microbial RPM increased considerably in 38 out of 45 pairs of samples having data from both mNGS with and without filter. For the BC positive samples, the effect of host depletion by filtration increased microbial target reads/million QC reads from 46 reads to 243 reads on average. A plot of the microbial RPM before and after filtration (Fig. 2a) showed that a wide range of enrichment (fold change in microbial RPM upon filtration compared to that without filtration) occurred amongst the samples. Fold-enrichment ranged from less than 1-log to more than 3-log (Fig. 2a, Supplementary Table 2). Particularly, the enrichment of microbial reads appears to be more effective with lower number of microbial reads entering the filter. In a small fraction of samples, enrichment was unsuccessful and number of reads declined slightly upon filtration. The reasons could be various and would be further discussed in the later session. In a fraction of samples, enrichment appeared insignificant, with no enrichment or even decrease in number of microbial reads upon filtration (samples 10, 12, 21, 22, 40, 48, 50). Interestingly, 3/7 of these were negative for BC1 and BC2, and 3/7 of these were negative for BC2, suggesting that there might be no significant level of pathogen content in these samples.

**Fig. 2.**
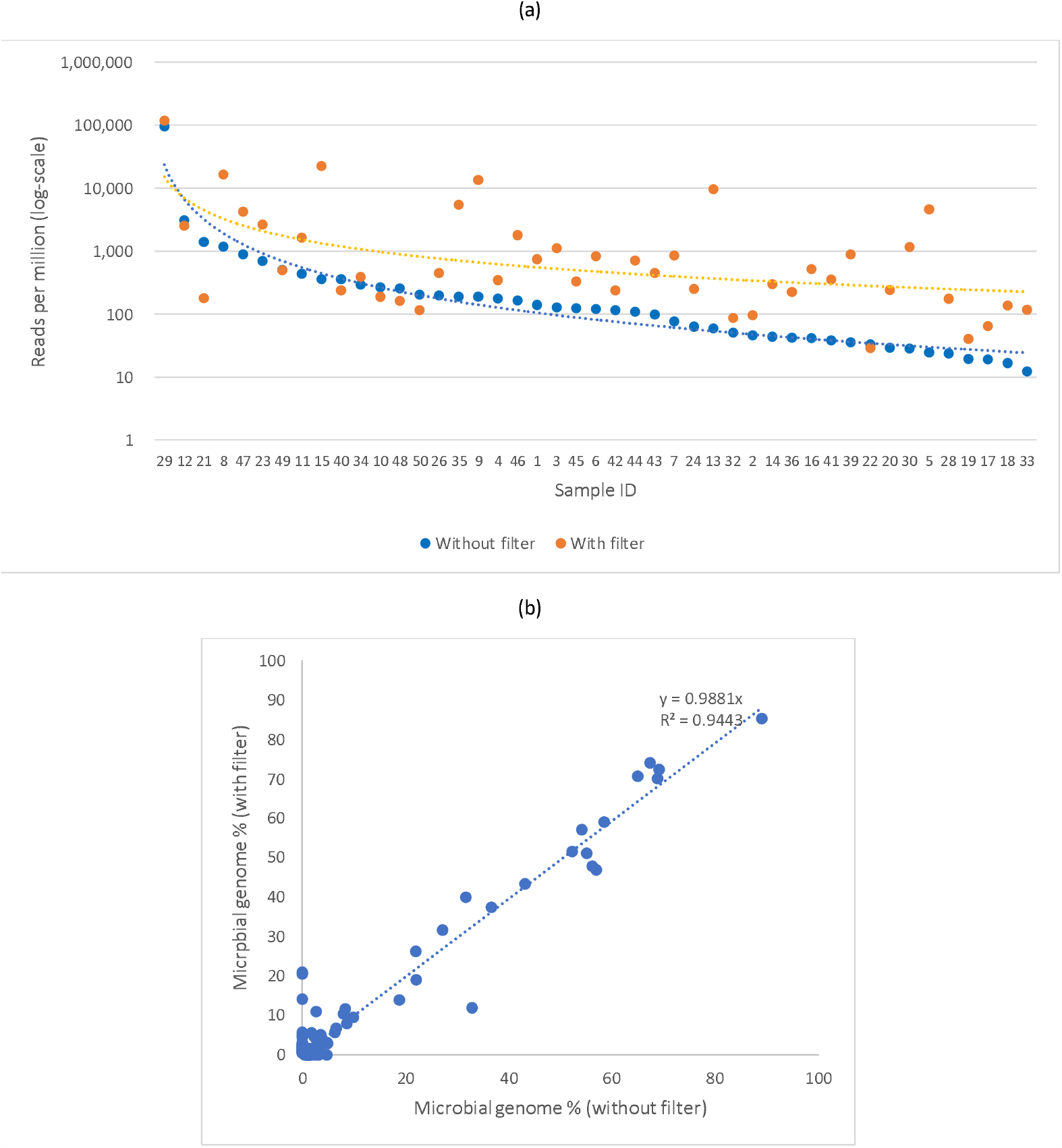
Comparisons of mNGS results from “With Filter” and “Without Filter” samples. (a) Comparison of sample reads (in Reads per Million) with and without filtration, arranged in order of decreasing reads for “Without Filter” samples. Y-axis (Sample output in RPM) is in log scale. (b) Plot of microbial genome % from “Without Filter” samples vs “With Filter” samples.

The % of individual species reads (microbial%) was computed by dividing each species reads to the total microbial reads (after removing human and unclassified reads). During infection, microbial% of the pathogen would be higher than that in normal individuals or a pre-defined baseline [16]. As such, we use the fold-enrichment of individual species microbial% in a sample over that in the NTC as one of the criteria to identify potential positive pathogens. Hence, it is important that the process of Devin filtration should not randomly alter the microbial% for each microorganism in a sample. To verify this, the microbial % reads for each microorganism in all samples with filter was correlated with that from the same samples without filter (Fig. 2b). Results showed that there was a tight correlation (R ^2^=0.96) in microbial % reads with and without filtration, and the correlation was close to 1:1. Hence, the process of Devin filtration does not affect proportion of microbial species in general.

### Comparison with cfDNA detection

Although this study was based on the use of gDNA for pathogen detection, a number of mNGS studies employs cfDNA as the target for pathogen detection. The strength and weakness of a cfDNA approach is discussed below. To investigate the correlation between gDNA and cfDNA approaches, cfDNA was harvested from the plasma from selected BC positive samples and subject to mNGS. Results showed that most of the expected pathogens were also detectable by cfDNA (Table 3). In the absence of filtration, the microbial RPM was similar between gDNA and cfDNA which was 280 and 140, respectively. However, with the use of filter, gDNA-based mNGS obtained significantly increased microbial RPM of an average of 2,359, while cfDNA-based method only achieved and average microbial RPM of 95 by filter, which was even slightly less than the process without filter. Hence, gDNA but not cfDNA was amenable to host-depletion and enrichment in microbial sequences. This suggests that signal derived from cfDNA would be closer to baseline noise, imposing a greater tendency for calling false positives.

**Table 3:** Comparison between genomic DNA-based mNGS and cell-free DNA-based mNGS in selected samples.

| No. | BC Results | Filter | Plasma cfDNA mNGS (Reads/Million QC Reads) |  |  |  |  |  | Pellet gDNA mNGS (Reads/Million QC Reads) |  |  |  |  |  |  |
| --- | --- | --- | --- | --- | --- | --- | --- | --- | --- | --- | --- | --- | --- | --- | --- |
|  |  |  | QC reads | A_halo | I_halo | Human | Microbial | Target | QC reads | A_halo | I_halo | Human | Microbial | Unclassified | Target |
| 9 | *Escherichia coli* | + | 17,271,416 | 0 | 1 | 678,789 | 33 | 10 | 26,129,510 | 5,497 | 4,671 | 695,456 | 13,515 | 280,860 | 106 |
|  |  | - | 19,409,969 | 1 | 1 | 773,167 | 28 | 9 | 24,721,756 | 520 | 902 | 851,295 | 189 | 147,094 | 3 |
| 11 | *Proteus mirabilis* | + | 18,717,399 | 2 | 5 | 891,739 | 284 | 0 | 38,070,697 | 5,376 | 7,883 | 768,171 | 1,645 | 216,924 | 159 |
|  |  | - | 17,581,154 | 6 | 14 | 895,122 | 501 | 0 | 49,385,166 | 1,556 | 3,195 | 1,595,623 | 871 | 396,393 | 41 |
| 14 | *Klebsiella pneumoniae* | + | 17,965,891 | 0 | 0 | 874,166 | 121 | 102 | 48,456,495 | 281 | 355 | 555,658 | 301 | 443,406 | 161 |
|  |  | - | 14,634,038 | 0 | 0 | 706,821 | 60 | 45 | 27,302,004 | 2 | 1 | 894,517 | 48 | 209,804 | 25 |
| 17 | MRSA | + | 10,598,680 | 0 | 0 | 50,970 | 49 | 0 | 36,865,299 | 284 | 312 | 596,557 | 65 | 402,782 | 1 |
|  |  | - | 15,827,810 | 0 | 0 | 931,437 | 12 | 1 | 35,954,996 | 66 | 9 | 1,137,207 | 28 | 317,077 | 1 |
| 19 | *Pseudomonas aeruginosa* | + | 20,592,770 | 0 | 0 | 54,732 | 109 | 2 | 26,843,560 | 441 | 488 | 781,736 | 42 | 217,293 | 13 |
|  |  | - | 20,332,529 | 0 | 1 | 154,006 | 201 | 4 | 27,897,183 | 34 | 25 | 841,496 | 20 | 158,426 | 2 |
| 23 | *Achromobacter species* | + | 17,092,585 | 2 | 4 | 776,858 | 99 | 79 | 30,817,981 | 22,533 | 15,655 | 577,856 | 3,257 | 380,699 | 325 |
|  |  | - | 14,707,563 | 0 | 0 | 55,473 | 65 | 2 | 36,357,904 | 2,380 | 2,515 | 755,295 | 697 | 239,112 | 229 |
| 30 | *Escherichia coli* | + | 19,799,176 | 0 | 0 | 30,474 | 89 | 7 | 27,963,103 | 754 | 1,377 | 889,600 | 1,453 | 106,816 | 93 |
|  |  | - | 20,271,797 | 1 | 3 | 378,187 | 342 | 8 | 26,037,075 | 29 | 60 | 822,646 | 28 | 177,236 | 1 |
| 43 | *Escherichia coli* | + | 12,236,113 | 0 | 0 | 80,867 | 43 | 3 | 28,916,267 | 1,467 | 2,810 | 705,206 | 453 | 290,064 | 28 |
|  |  | - | 12,250,956 | 0 | 0 | 8,910 | 36 | 3 | 33,734,655 | 367 | 682 | 1,102,967 | 135 | 260,423 | 5 |
| 49 | *Escherichia coli* | + | 12,066,105 | 0 | 0 | 34,571 | 26 | 2 | 23,940,664 | 913 | 1,248 | 795,679 | 499 | 201,662 | 25 |
|  |  | - | 14,780,596 | 0 | 2 | 831,639 | 34 | 13 | 24,586,630 | 350 | 697 | 783,625 | 505 | 209,357 | 39 |

## DISCUSSION

### Enhancement of mNGS sensitivity by host depletion

One long-standing challenge of mNGS is the overwhelming presence of host DNA background in human biospecimens especially the whole blood. Although different host depletion methods have been tested, they are not practical in the clinical workflow in addition to widely varied host depletion effect. The host cell/DNA depletion by filtration tested in this study showed significant level of enrichment of microbial reads comparing to the sample processed without filter, especially for the samples with low microbial content. The host depletion indeed enhanced the sensitivity from 63.6% for mNGS without filter to 81.8% to mNGS with filter, suggesting that filtration can enrich microbial content in the gDNA portion of the sample using the process tested in this study. On the other hand, cfDNA was compared to this gDNA based mNGS approached in this study because most of the reported studies were based on cfDNA. No significant enrichment of microbial content in the cfDNA portion was observed between samples with and without filtration. Thus, we demonstrated in this study that gDNA based mNGS was amenable to host-depletion and enrichment in microbial sequences by filtration. With the easy process of filtration together with 100bp sequencing and analysis, it is possible now to establish a clinical applicable workflow can be completed within 24 hours with enhanced sensitivity comparing to other mNGS methods.

### Criteria for pathogen identification by mNGS

Another major challenge of mNGS is that currently it lacks consensus of the definitive criteria for distinguishing positive pathogen from background. It’s documented that most of the reagents used for mNGS have been found to also introduce foreign DNA during the sequencing process. This phenomenon has been described as “kit-ome”, which will seriously confound the sample result [8]. Therefore, it is important to capture the nucleic acid background of the environment and reagents used for sequencing, which can help filter out contaminated background readings during the interpretation of the results. In this study, a template-free control (NTC) was used in the mNGS analysis to set the baseline for making positive calls. We used the tentative cut-off criteria fold change (microbe %_SAMPLE_ : microbe %_NTC_) ≥ 5 and microbial RPM ≥ 5 for potential pathogen identification to achieve sensitivity as well as remove potential false positive from the background as much as possible. However, the current criteria may miss some potential positive. For expample, in sample 33 of which the urine culture is positive for *Aerococcus urinae*, this species showed significant fold change of microbe %_SAMPLE_ : microbe %_NTC_) but microbial RPM of 3.18 didn’t pass this species as positive. Further refinement of such criteria need to be tested in a much larger sample size. On the other hand, in some of the samples especially those processed at the same time, there were some identified positives may contributed by the process background such as sample 3 and 4, sample 7 and 8, sample 13 and 15. Further refinement of this would be 1) to apply NTC to every batch of sample processing and sequencing; 2) to build a database of all NTCs of the same batch of reagents.

### Clinical accuracy and diagnostic yield

Karius offers a commercial cfDNA based mNGS laboratory developed test (LDT) for sepsis diagnosis. The test received CLIA and New York State approvals, with a claimed sensitivity of 92.9% and a claimed specificity of 62.7%. Working on pathogen detection in cerebrospinal fluid (CSF) by mNGS, Miller et al [16] reported a claimed sensitivity of 73% and a claimed specificity of 99%. In some study [17], sensitivity and specificity were not explicitly stated. In this study, a sensitivity of 81.8% was reported.

Karius reported 2.7x more causal pathogens than initial blood culture and 1.3x more causal pathogens than all microbiology tests. Cheng and Yu [4] reported in their study that 77 (86%) were positive for 1 or more pathogens using NGS, compared to 50 (56%) which were positive using traditional detection methods. In this study, the diagnostic yield by mNGS with host depletion by filtration (90%) was significantly higher than clinical microbial culture (56%). Hence, mNGS was able to provide more positive results than conventional pathogen detection.

A previous study showed that the rate of positive mNGS results was constant over the different time points after sepsis developed, while the positivity of blood culture decreased at later time points [10]. In another study, cell-free DNA sequencing could still identify fungus such as *P. jirovecii* or *Aspergillus* species among patients receiving effective antifungal agents [18]. In this study, the second blood culture done after antibiotic treatment had significantly lower sensitivity compared to mNGS, and there were several cases where an organism is detected consistent with BC1 but was negative by BC2. Hence, our findings were consistent with other studies in that mNGS possessed greater sensitivity for those who had blood sampled prior to effective antimicrobial agent exposure, suggesting mNGS could be a more effective detection method for patients undergoing antibiotic treatment.

### Future prospects and concluding remarks

The target independent identification of potential pathogen by mNGS has made it a very promising tools for clinical microbiology comparing to other molecular tests. It can detect different microorganisms simultaneously, for example, *Escherichia coli* and *Candida albicans* in sample 43. It can detect microorganisms unculturable by routine culture method, for example, Torque teno virus in sample 41. In conclusion, mNGS was able to [19] identify majority of the organisms detected by microbial culture and showed a good correlation with routine clinical results. In addition, it provided more results, potentially able to assuage the short-coming of poor diagnostic yield of conventional culture. However, the demonstration of clinical utility is required for such test to be applied for clinical diagnostics. Stanford Healthcare set out to determine the real-world clinical utility of the Karius test in a subset of immunocompromised patients [20], with the conclusion that, in 48 cases, positive impact was observed in 6 (7.3%) cases, negative impact observed in 3 (3.7%) cases, and no impact in 71 (86.6%) cases. More studies need to be performed to assess clinical utility in different group of patients.

The other important questions to consider are whether mNGS has the diagnostic accuracy required to complement currently available diagnostics, and how best to integrate mNGS into current testing algorithms. One possibility is to reserve mNGS as last resort for cases for which conventional microbiological testing has failed to provide an answer, or to include mNGS more proximally in testing in parallel to conventional tests. These are currently an active subject of discussion in the microbiology field that needs to be determined empirically.

## Data Availability

All data produced in the present work are contained in the manuscript

---

**Supplemental Table 1.** Comparison of target reads from analyses based on 100bp, 120bp and 150bp read lengths.

| No | BC Results<br>(BC1+BC2+) | Length | mNGS w/ filter (Reads/Million QC Reads) |  |  |  |  |  |  | mNGS w/o filter (Reads/Million QC Reads) |  |  |  |  |  |  |
| --- | --- | --- | --- | --- | --- | --- | --- | --- | --- | --- | --- | --- | --- | --- | --- | --- |
|  |  |  | QC reads | A_halo | I_halo | Human | Microbial | Unclassified | Target | QC reads | A_halo | I_halo | Human | Microbial | Unclassified | Target |
| 5 | Enterococcus faecalis | 150bp | 29,415,977 | 53,454 | 11,599 | 472,560 | 884 | 461,503 | 762 | 33,552,849 | 10,817 | 3,886 | 769,101 | 128 | 216,069 | 82 |
|  |  | 120bp | 29,608,326 | 53,159 | 11,538 | 468,783 | 873 | 465,647 | 754 | 33,917,148 | 10,755 | 3,865 | 762,573 | 123 | 222,684 | 81 |
|  |  | 100bp | 30,624,819 | 51,268 | 11,119 | 448,627 | 835 | 488,151 | 720 | 33,650,802 | 10,820 | 3,885 | 747,721 | 121 | 237,454 | 81 |
| 9 | Escherichia coli | 150bp | 25,939,257 | 5,614 | 4,732 | 717,892 | 16,798 | 254,963 | 116 | 24,539,226 | 525 | 910 | 880,994 | 240 | 117,331 | 3 |
|  |  | 120bp | 26,055,023 | 5,555 | 4,708 | 711,711 | 15,364 | 262,661 | 115 | 24,648,590 | 523 | 907 | 875,436 | 216 | 122,919 | 3 |
|  |  | 100bp | 26,129,510 | 5,497 | 4,671 | 695,456 | 13,515 | 280,860 | 106 | 24,721,756 | 520 | 902 | 851,295 | 189 | 147,094 | 3 |
| 10 | Proteus mirabilis | 150bp | 37,795,374 | 273 | 421 | 500,427 | 202 | 498,676 | 77 | 21,311,225 | 34 | 47 | 746,414 | 258 | 121,703 | 105 |
|  |  | 120bp | 37,990,451 | 272 | 420 | 496,703 | 195 | 502,410 | 77 | 21,406,413 | 34 | 47 | 740,732 | 246 | 127,406 | 104 |
|  |  | 100bp | 38,146,610 | 270 | 416 | 488,233 | 189 | 510,892 | 76 | 21,471,857 | 34 | 47 | 719,489 | 232 | 148,740 | 104 |
| 11 | Proteus mirabilis | 150bp | 37,801,114 | 5,446 | 7,981 | 798,519 | 2,076 | 185,978 | 163 | 49,097,239 | 1,571 | 3,225 | 1,655,603 | 1,095 | 339,272 | 41 |
|  |  | 120bp | 37,964,488 | 5,418 | 7,949 | 790,765 | 1,880 | 193,988 | 162 | 49,273,381 | 1,564 | 3,213 | 1,641,589 | 994 | 351,674 | 41 |
|  |  | 100bp | 38,070,697 | 5,376 | 7,883 | 768,171 | 1,645 | 216,924 | 159 | 49,385,166 | 1,556 | 3,195 | 1,595,623 | 871 | 396,393 | 41 |
| 14 | Klebsiella pneumoniae | 150bp | 47,771,606 | 286 | 362 | 580,520 | 312 | 418,520 | 168 | 27,124,936 | 2 | 1 | 925,780 | 50 | 179,538 | 26 |
|  |  | 120bp | 48,173,290 | 284 | 359 | 572,212 | 306 | 426,839 | 164 | 27,232,493 | 2 | 1 | 918,524 | 47 | 186,256 | 26 |
|  |  | 100bp | 48,456,495 | 281 | 355 | 555,658 | 301 | 443,406 | 161 | 27,302,004 | 2 | 1 | 894,517 | 48 | 209,804 | 25 |
| 17 | MRSA | 150bp | 36,397,462 | 289 | 317 | 618,042 | 76 | 381,275 | 1 | 35,652,507 | 66 | 9 | 1,175,811 | 26 | 276,966 | 1 |
|  |  | 120bp | 36,671,574 | 287 | 315 | 611,499 | 70 | 387,829 | 1 | 35,835,593 | 66 | 9 | 1,168,009 | 26 | 285,749 | 1 |
|  |  | 100bp | 36,865,299 | 284 | 312 | 596,557 | 65 | 402,782 | 1 | 35,954,996 | 66 | 9 | 1,137,207 | 28 | 317,077 | 1 |
| 42 | Escherichia coli | 150bp | 23,900,937 | 134 | 29 | 874,048 | 308 | 125,480 | 7 | 27,046,543 | 848 | 997 | 960,831 | 150 | 139,349 | 5 |
|  |  | 120bp | 24,088,163 | 133 | 29 | 865,630 | 280 | 133,927 | 6 | 27,046,543 | 845 | 992 | 956,568 | 149 | 138,731 | 5 |
|  |  | 100bp | 24,212,385 | 132 | 29 | 839,711 | 240 | 159,888 | 6 | 27,332,019 | 842 | 988 | 929,766 | 128 | 173,861 | 5 |
| 43 | Escherichia coli & Candida albicans | 150bp | 28,522,975 | 1,491 | 2,860 | 738,941 | 559 | 256,148 | 30 | 33,366,587 | 370 | 690 | 1,144,729 | 153 | 213,782 | 5 |
|  |  | 120bp | 28,761,517 | 1,481 | 2,839 | 729,079 | 514 | 266,088 | 29 | 33,366,587 | 369 | 687 | 1,139,650 | 152 | 212,834 | 5 |
|  |  | 100bp | 28,916,267 | 1,467 | 2,810 | 705,206 | 453 | 290,064 | 28 | 33,734,655 | 367 | 682 | 1,102,967 | 135 | 260,423 | 5 |
| 45 | Klebsiella pneumoniae | 150bp | 18,920,723 | 144 | 19 | 748,746 | 355 | 250,737 | 247 | 27,717,908 | 71 | 145 | 959,664 | 140 | 169,515 | 108 |
|  |  | 120bp | 19,058,784 | 143 | 19 | 742,607 | 344 | 256,887 | 245 | 27,879,711 | 71 | 145 | 953,435 | 138 | 177,299 | 107 |
|  |  | 100bp | 19,151,162 | 142 | 19 | 723,721 | 332 | 275,785 | 242 | 27,980,656 | 70 | 143 | 927,259 | 141 | 204,210 | 106 |
| 46 | Escherichia coli | 150bp | 33,714,921 | 3,121 | 1,260 | 729,035 | 1,872 | 264,711 | 1,246 | 21,397,594 | 29 | 17 | 750,749 | 144 | 121,037 | 100 |
|  |  | 120bp | 33,998,045 | 3,101 | 1,252 | 720,682 | 1,841 | 273,124 | 1,210 | 21,560,856 | 28 | 17 | 747,034 | 144 | 127,506 | 94 |
|  |  | 100bp | 34,189,349 | 3,073 | 1,240 | 697,513 | 1,798 | 296,376 | 1,149 | 21,665,310 | 28 | 17 | 728,305 | 144 | 147,872 | 92 |
| 49 | Escherichia coli | 150bp | 23,784,973 | 926 | 1,265 | 828,916 | 657 | 168,236 | 27 | 24,406,342 | 356 | 707 | 830,553 | 623 | 162,346 | 42 |
|  |  | 120bp | 23,880,506 | 920 | 1,259 | 820,887 | 583 | 176,352 | 26 | 24,516,514 | 353 | 703 | 818,939 | 569 | 174,077 | 40 |
|  |  | 100bp | 23,940,664 | 913 | 1,248 | 795,679 | 499 | 201,662 | 25 | 24,586,630 | 350 | 697 | 783,625 | 505 | 209,357 | 39 |

**Supplemental Table 2.** Sample reads from all samples.

| No. | BC1 Results | BC1 | BC2 | mNGS w/ filter (Reads/Million QC Reads) |  |  |  |  |  | mNGS w/o filter (Reads/Million QC Reads) |  |  |  |  |  |
| --- | --- | --- | --- | --- | --- | --- | --- | --- | --- | --- | --- | --- | --- | --- | --- |
|  |  |  |  | QC reads | A_halo | I_halo | Human | Microbial | Unclassified | QC reads | A_halo | I_halo | Human | Microbial | Unclassified |
| 1 | Staphylococcus aureus (MRSA) | + | - | 26,874,647 | 284,561 | 272,359 | 258,754 | 746 | 183,579 | 43,224,176 | 30,098 | 42,845 | 739,796 | 140 | 187,120 |
| 2 | negative | - | - | 26,186,906 | 11,233 | 10,953 | 691,594 | 96 | 286,124 | 28,118,220 | 4,705 | 3,987 | 757,768 | 46 | 233,494 |
| 3 | negative | - | - | 27,738,717 | 215,429 | 273,669 | 308,884 | 1,112 | 200,906 | 23,447,427 | 41,580 | 53,692 | 719,329 | 129 | 185,270 |
| 4 | negative | - | - | 28,547,723 | 103,619 | 224,762 | 343,899 | 348 | 327,372 | 29,211,386 | 21,550 | 49,031 | 597,688 | 177 | 331,554 |
| 5 | Proteus mirabilis | + | - | 27,883,257 | 247,432 | 294,721 | 312,346 | 4,639 | 140,862 | 32,394,909 | 1,018 | 759 | 837,179 | 25 | 161,019 |
| 6 | Enterococcus faecalis | + | + | 30,624,819 | 51,268 | 11,119 | 448,627 | 835 | 488,151 | 33,650,802 | 10,820 | 3,885 | 747,721 | 121 | 237,454 |
| 7 | negative | - | - | 23,902,590 | 2,463 | 2,324 | 930,937 | 852 | 63,425 | 33,735,056 | 109 | 120 | 852,806 | 77 | 146,888 |
| 8 | negative | - | - | 4,584,817 | 9,610 | 17,023 | 877,372 | 16,560 | 79,435 | 26,743,891 | 313 | 313 | 901,470 | 1,178 | 96,726 |
| 9 | E. coli | + | + | 26,129,510 | 5,497 | 4,671 | 695,456 | 13,515 | 280,860 | 24,721,756 | 520 | 902 | 851,295 | 189 | 147,094 |
| 10 | Proteus mirabilis | + | + | 38,146,610 | 270 | 416 | 488,233 | 189 | 510,892 | 21,471,857 | 39 | 54 | 828,388 | 267 | 171,252 |
| 11 | Proteus mirabilis | + | + | 38,070,697 | 5,376 | 7,883 | 768,171 | 1,645 | 216,924 | 49,385,166 | 779 | 1,600 | 798,754 | 436 | 198,431 |
| 12 | Klebsiella pneumoniae | + | - | 38,659,564 | 63 | 76 | 619,828 | 2,529 | 377,504 | 34,975,786 | 139 | 282 | 687,604 | 3,111 | 308,864 |
| 13 | negative | - | - | 25,353,467 | 8,470 | 8,386 | 763,949 | 9,690 | 209,505 | 25,368,316 | 53 | 44 | 847,812 | 60 | 152,031 |
| 14 | Klebsiella pneumoniae | + | + | 48,456,495 | 281 | 355 | 555,658 | 301 | 443,406 | 27,302,004 | 2 | 0 | 809,979 | 44 | 189,975 |
| 15 | negative | - | - | 25,631,842 | 13,736 | 14,578 | 683,575 | 22,436 | 265,675 | 37,609,174 | 632 | 648 | 824,551 | 360 | 173,810 |
| 16 | negative | - | - | 31,733,417 | 2,568 | 2,046 | 721,508 | 520 | 273,358 | 39,535,385 | 157 | 154 | 821,625 | 42 | 178,022 |
| 17 | MRSA | + | + | 36,865,299 | 284 | 312 | 596,557 | 65 | 402,782 | 35,954,996 | 46 | 6 | 781,915 | 19 | 218,014 |
| 18 | Proteus mirabilis | + | - | 28,052,179 | 824 | 1,198 | 702,840 | 137 | 295,000 | 27,209,478 | 16 | 22 | 847,032 | 17 | 152,913 |
| 19 | Pseudomonas aeruginosa | + | - | 27,246,168 | 436 | 482 | 754,048 | 40 | 244,994 | 27,897,183 | 34 | 25 | 841,496 | 20 | 158,426 |
| 20 | negative | - | - | 29,232,228 | 452 | 826 | 539,168 | 243 | 459,310 | 26,756,735 | 141 | 146 | 855,335 | 29 | 144,348 |
| 21 | Escherichia coli | + | - | 20,165,261 | 494 | 725 | 574,594 | 180 | 424,008 | 22,092,098 | 166 | 435 | 740,207 | 1,407 | 257,786 |
| 22 | Klebsiella pneumoniae | + | - | 39,491,616 | 248 | 182 | 859,433 | 29 | 140,108 | 33,406,603 | 90 | 148 | 870,375 | 33 | 129,353 |
| 23 | Achromobacter species | + | - | 31,387,305 | 22,129 | 15,356 | 556,667 | 2,641 | 403,207 | 36,357,904 | 2,380 | 2,515 | 755,295 | 697 | 239,112 |
| 24 | negative | - | - | 35,345,932 | 469 | 488 | 810,182 | 251 | 188,609 | 55,344,131 | 113 | 80 | 874,802 | 63 | 124,942 |
| 25 | negative | - | - |  |  |  |  |  |  |  |  |  |  |  |  |
| 26 | negative | - | - | 34,689,495 | 193 | 307 | 770,246 | 449 | 228,805 | 15,497,865 | 61 | 55 | 141,362 | 199 | 858,324 |
| 27 | negative | - | - | 28,608,412 | 23,675 | 20,143 | 748,024 | 8,028 | 200,130 |  |  |  |  |  |  |
| 28 | negative | - | - | 27,280,309 | 791 | 1,072 | 786,959 | 176 | 211,002 | 30,130,081 | 16 | 14 | 845,505 | 24 | 154,441 |
| 29 | Pseudomonas aeruginosa | + | - | 28,857,811 | 73,190 | 48,709 | 559,774 | 117,810 | 200,517 | 29,346,400 | 18,548 | 25,655 | 744,089 | 95,734 | 115,974 |
| 30 | Escherichia coli | + | - | 28,158,790 | 745 | 1,362 | 855,347 | 1,166 | 141,380 | 26,037,075 | 29 | 60 | 822,646 | 28 | 177,236 |
| 31 | Proteus mirabilis; Clostridium perfringens | + | - |  |  |  |  |  |  |  |  |  |  |  |  |
| 32 | negative | - | - | 29,656,175 | 60 | 48 | 676,036 | 87 | 323,769 | 29,328,723 | 116 | 100 | 863,037 | 51 | 136,696 |
| 33 | negative | - | - | 32,398,145 | 49 | 2 | 826,248 | 118 | 173,582 | 29,586,937 | 9 | 2 | 855,433 | 12 | 144,544 |
| 34 | negative | - | - | 29,975,987 | 2,338 | 1,747 | 777,967 | 390 | 217,557 | 19,380,173 | 235 | 200 | 854,605 | 294 | 144,666 |
| 35 | negative | - | - | 30,750,148 | 23,912 | 18,841 | 690,882 | 5,450 | 260,914 | 33,075,784 | 530 | 376 | 860,051 | 190 | 138,853 |
| 36 | Escherichia coli | + | - | 32,270,957 | 694 | 716 | 783,895 | 225 | 214,470 | 21,735,684 | 52 | 37 | 873,070 | 42 | 126,799 |
| 37 | negative | - | - |  |  |  |  |  |  |  |  |  |  |  |  |
| 38 | Escherichia coli | + | - | 19,794,833 | 373 | 537 | 649,344 | 1,740 | 348,006 |  |  |  |  |  |  |
| 39 | negative | - | - | 25,584,865 | 761 | 669 | 843,909 | 887 | 153,774 | 26,214,472 | 58 | 45 | 858,255 | 36 | 141,606 |
| 40 | negative | - | - | 30,626,048 | 1,911 | 1,558 | 669,674 | 240 | 326,617 | 30,511,094 | 858 | 903 | 729,191 | 358 | 268,691 |
| 41 | negative | - | - | 25,914,287 | 359 | 120 | 876,129 | 356 | 123,037 | 32,895,717 | 93 | 137 | 857,240 | 39 | 142,493 |
| 42 | Escherichia coli | + | + | 24,212,385 | 132 | 29 | 839,711 | 240 | 159,888 | 27,332,019 | 761 | 894 | 840,972 | 116 | 157,257 |
| 43 | Escherichia coli & Candida albicans | + | + | 28,916,267 | 1,467 | 2,810 | 705,206 | 453 | 290,064 | 33,734,655 | 269 | 500 | 808,287 | 99 | 190,846 |
| 44 | Klebsiella pneumoniae | + | - | 28,530,594 | 3,477 | 2,916 | 588,260 | 714 | 404,633 | 27,115,403 | 248 | 244 | 785,449 | 109 | 213,950 |
| 45 | Klebsiella pneumoniae | + | + | 19,151,162 | 142 | 19 | 723,721 | 332 | 275,785 | 27,980,656 | 62 | 126 | 819,262 | 125 | 180,425 |
| 46 | Escherichia coli | + | + | 34,189,349 | 3,073 | 1,240 | 697,513 | 1,798 | 296,376 | 21,665,310 | 32 | 19 | 831,051 | 165 | 168,733 |
| 47 | negative | - | - | 22,696,014 | 3,126 | 4,740 | 715,638 | 4,254 | 272,243 | 26,658,180 | 1,198 | 1,488 | 798,436 | 893 | 197,986 |
| 48 | negative | - | - | 30,225,187 | 117 | 176 | 793,952 | 162 | 205,593 | 25,692,243 | 178 | 275 | 888,051 | 257 | 111,238 |
| 49 | Escherichia coli | + | + | 23,940,664 | 913 | 1,248 | 795,679 | 499 | 201,662 | 24,586,630 | 352 | 701 | 787,931 | 507 | 210,508 |
| 50 | negative | - | - | 22,923,655 | 25 | 31 | 596,744 | 117 | 403,083 | 27,452,004 | 237 | 486 | 700,539 | 204 | 298,533 |

**Supplemental Table 3.**
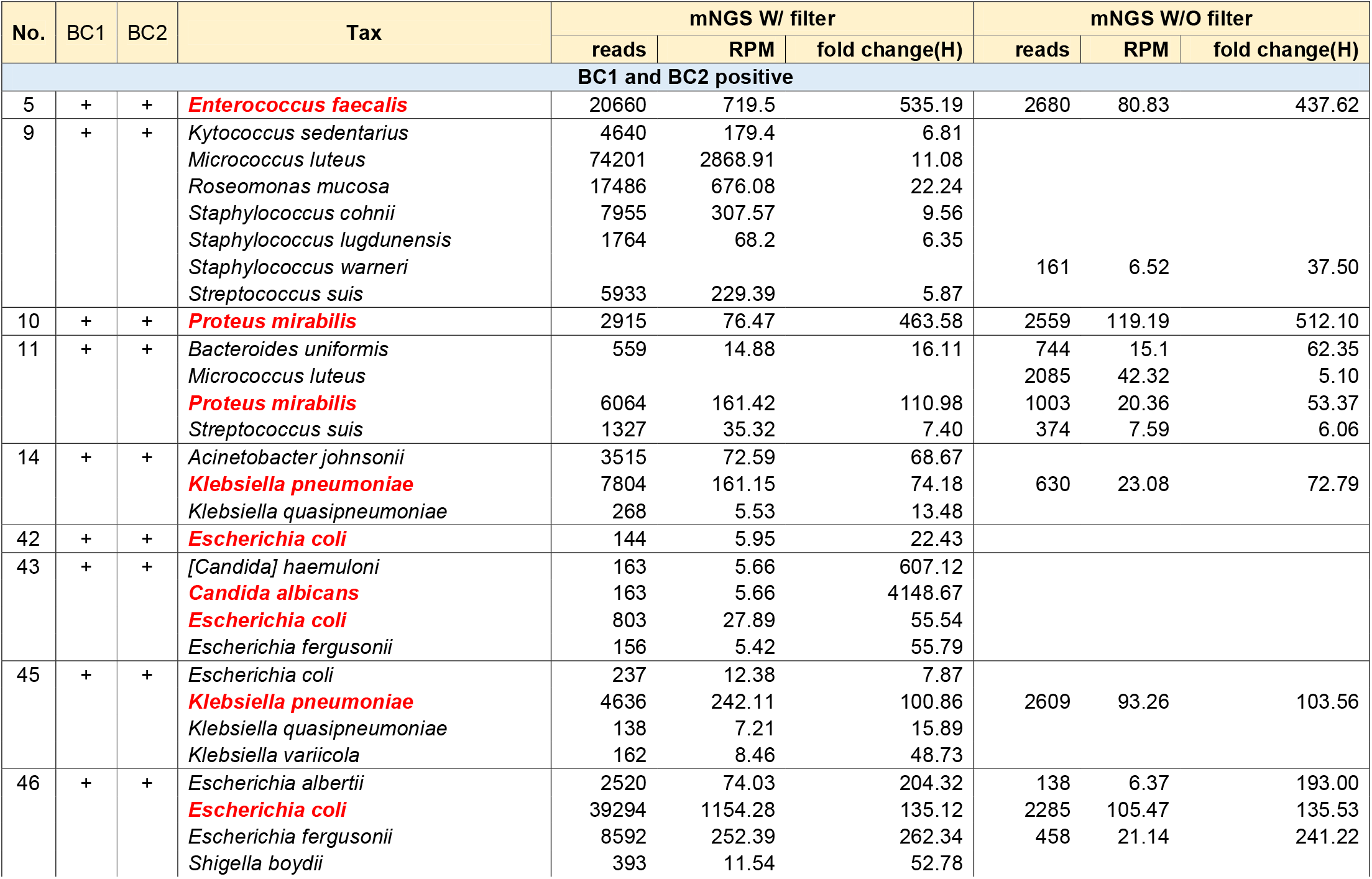
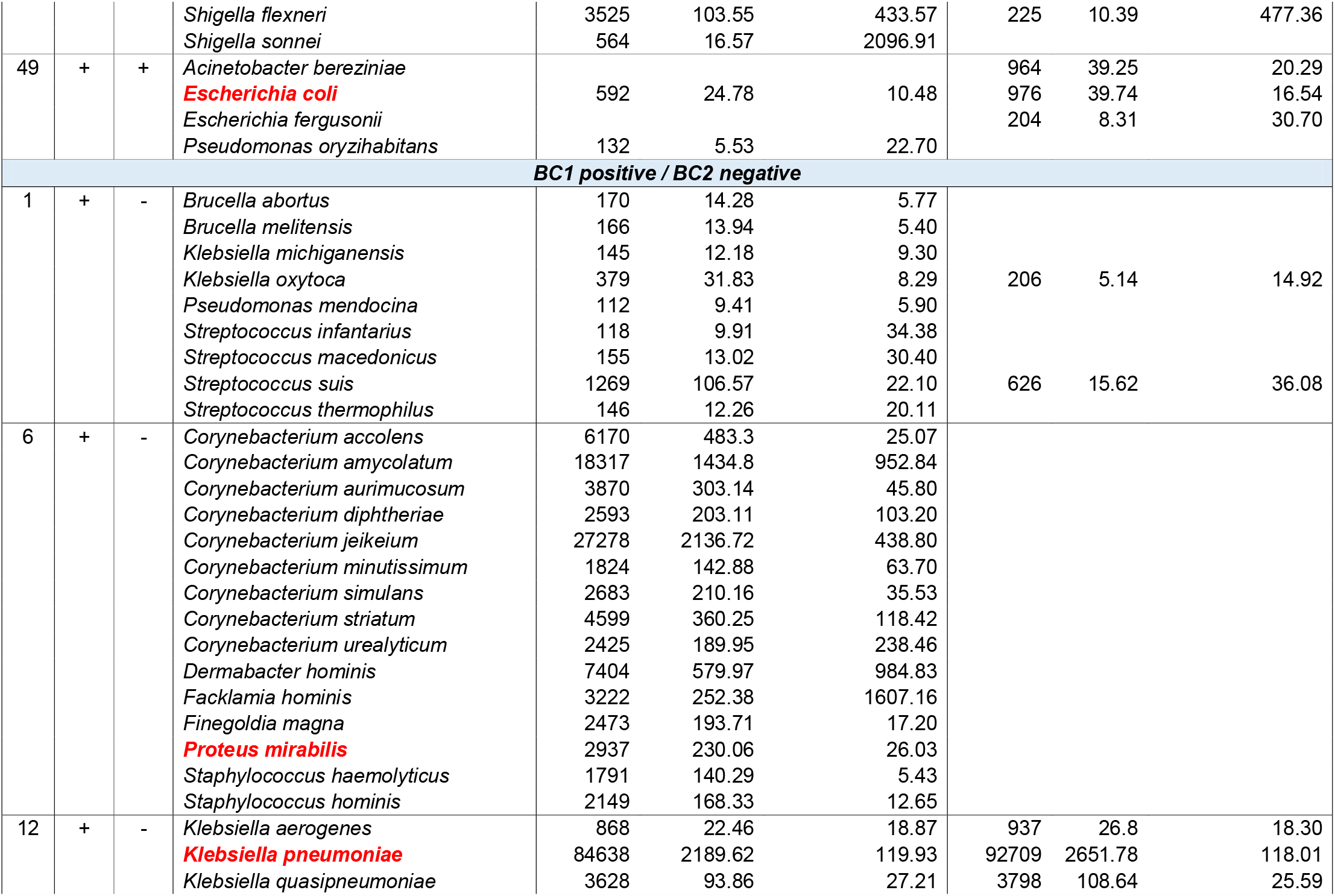
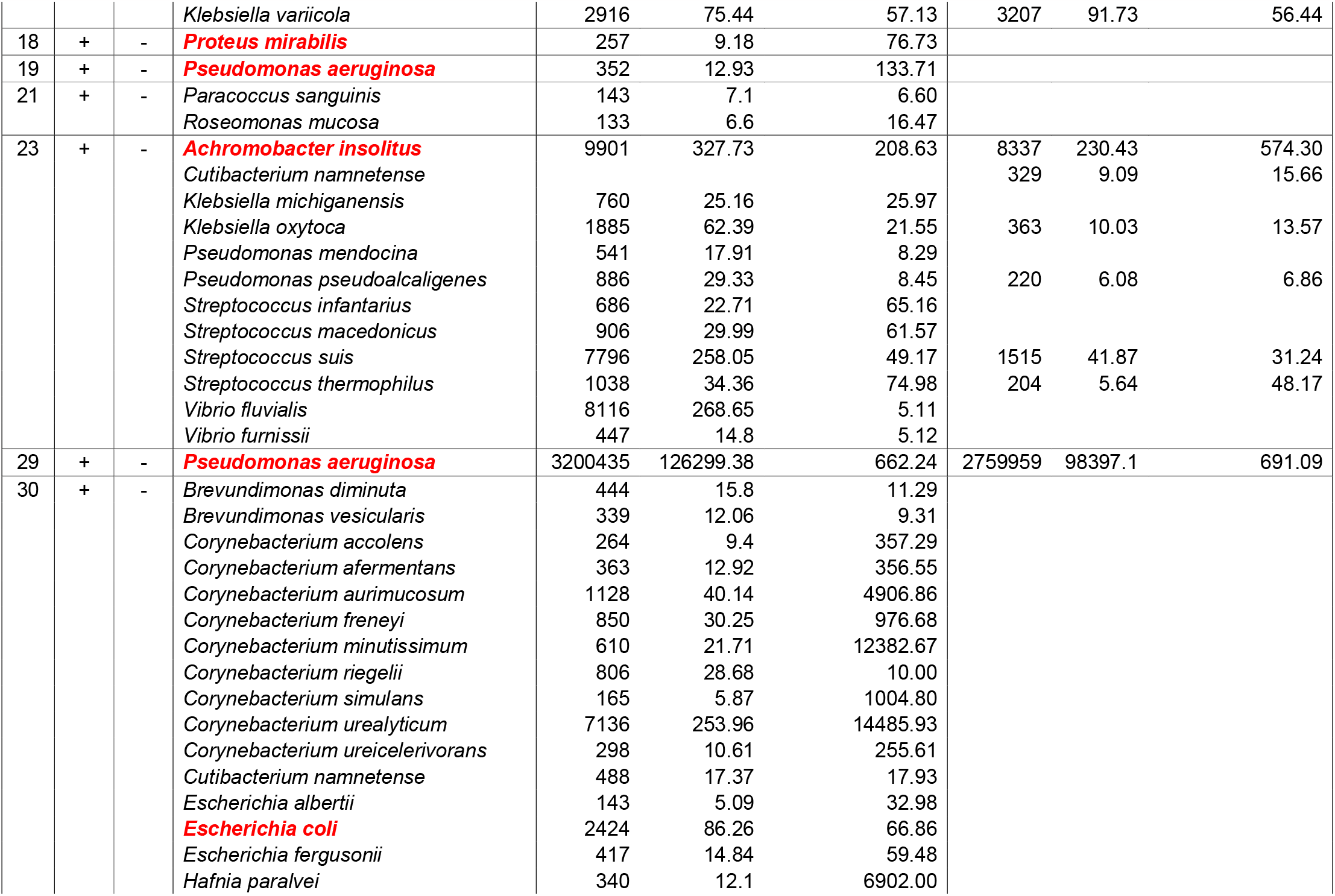
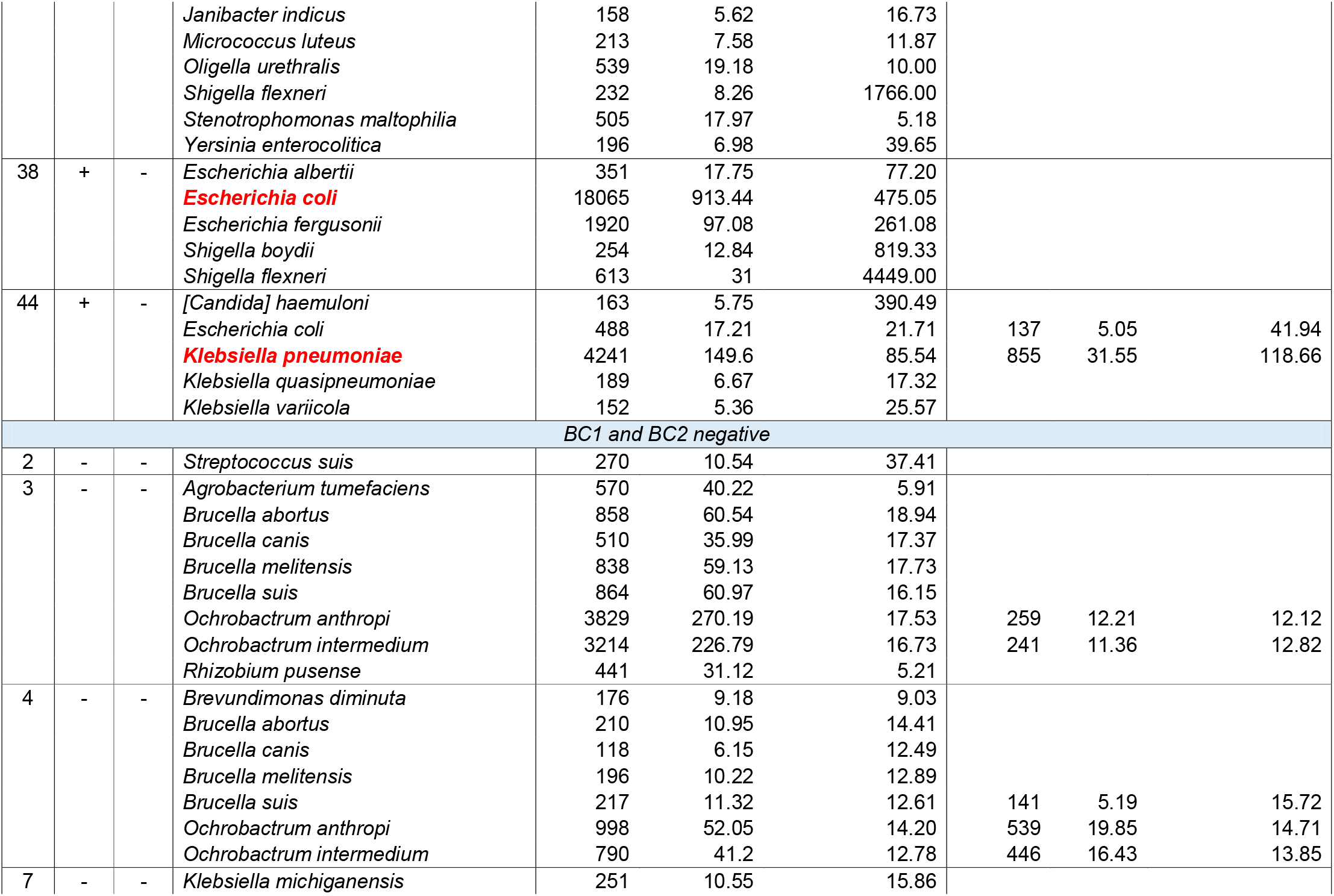
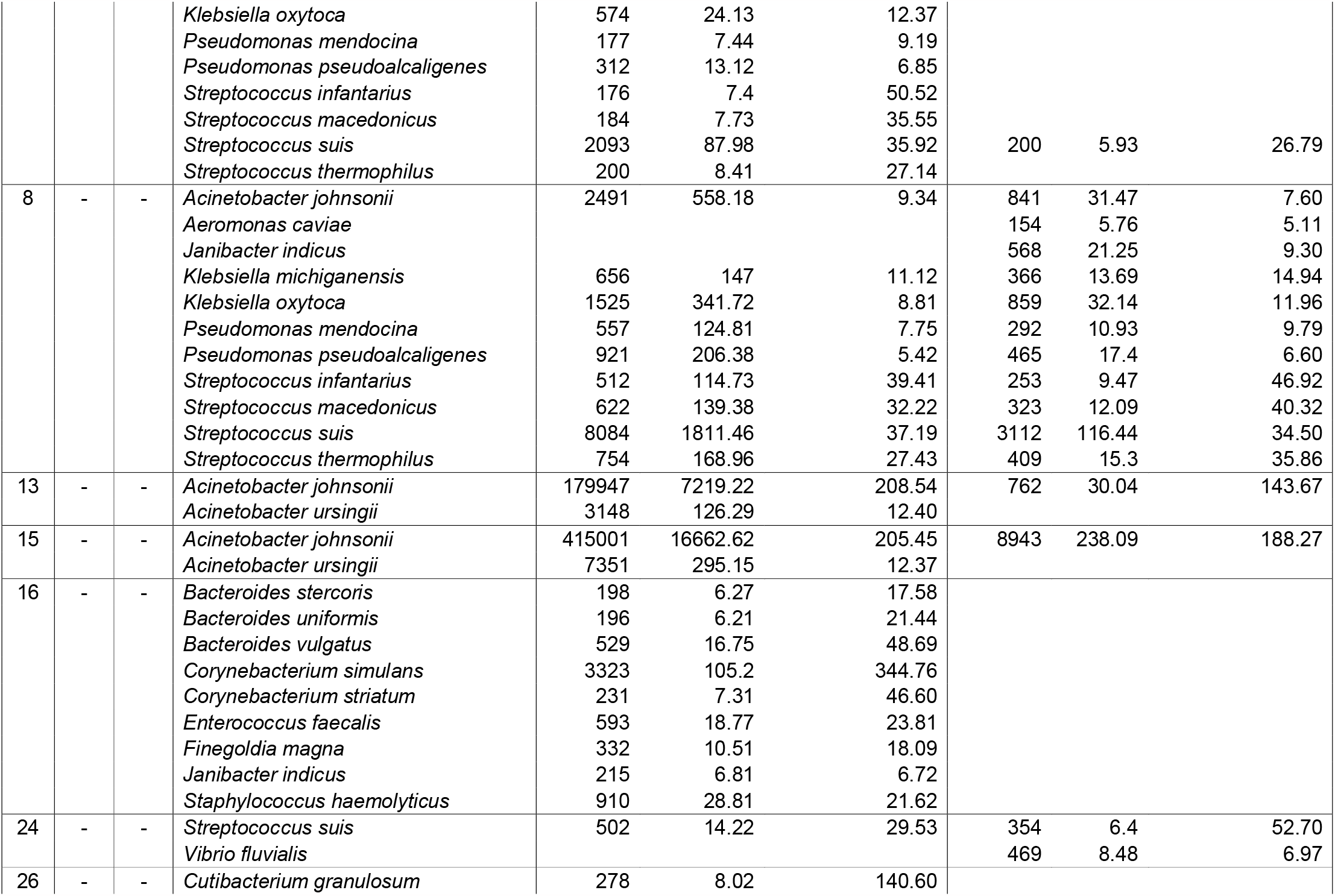
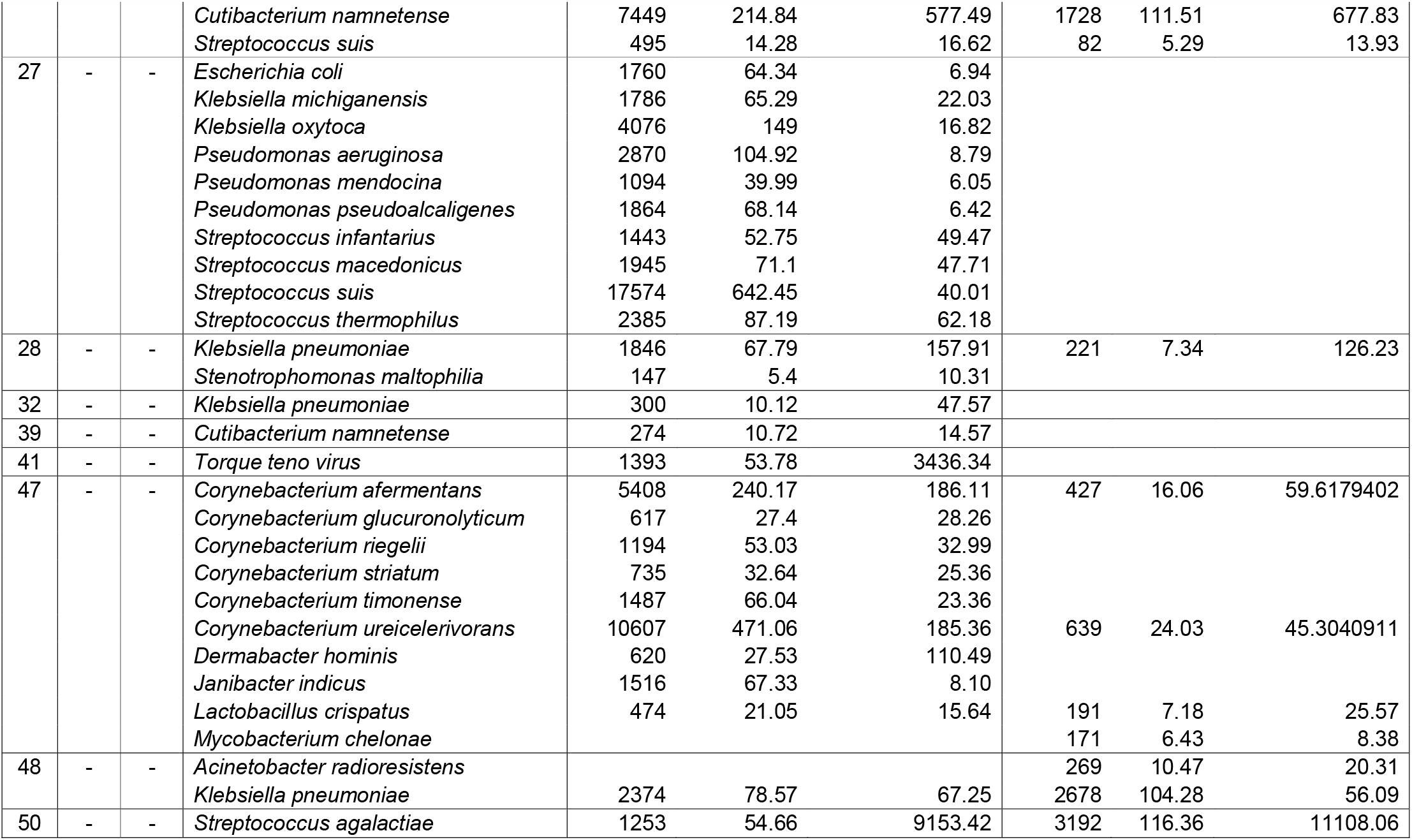
Complete list of potential pathogens Identified by mNGS (Fold change (% Microbialsample/% MicrobialNTC >=5 and RPM >=5)

